# Early detection of severe COVID-19 disease patterns define near real-time personalised care, bioseverity in males, and decelerating mortality rates

**DOI:** 10.1101/2020.05.08.20088393

**Authors:** Marcela P. Vizcaychipi, Claire L Shovlin, Michelle Hayes, Suveer Singh, Linsey Christie, Alice Sisson, Roger Davies, Christopher Lockie, Alice Howard, Alexander Brown, Alex McCarthy, Monica Popescu, Amandeep Gupta, James Armstrong, Hisham Said, Timothy Peters, Richard T Keays, ChelWest COVID-19 Consortium

## Abstract

**Background:** COVID-19 is a global health emergency. Recent data indicate a 50% mortality rate across UK intensive care units.

**Methods:** A single institution, two-centre retrospective analysis following implementation of a Decision Support tool and real-time data dashboard for early detection of patients requiring personalised enhanced care, focussing on respiratory rate, diastolic blood pressure, oxygenation indices, C-reactive protein, D-dimer and ferritin. Protocols differing from conventional practice included high-dose prophylactic anticoagulation for all COVID-19 positive patients and prescription of antioxidants.

**Results:** By 22/04/2020, 923 patients tested COVID-19 positive. 569 patients (61.7%) were male. The majority presented with advanced disease: interquartile ranges were C-reactive protein 44.9-179mg/L, D-dimer 1070-3802ng/mL, and ferritin 261-1208µg/L. Completed case fatality rates were 25.1% [95% CI 20.0, 30.0] in females, 40.5% [95% CI 35.9, 45.0] in males. 139 patients were admitted to intensive care where current death rates are 16.2% [95% CI 3.8, 28.7] in females, 38.2% [95% CI 28.6, 47.8] in males with no trends for differences based on ethnicity. A real-time traffic lights dashboard enabled rapid assessment of patients using critical parameters to accelerate adjustments to management protocols. In total 513 (55.6%) of patients were flagged as high risk for thromboembolic disease, exceeding the numbers flagged for respiratory deteriorations (N=391, 42.4%), or cytokine storm (N=68, 7.4%). There was minimal evidence that age was associated with disease severity, but males had higher levels of all dashboard indices, particularly C-reactive protein and ferritin (p<0.0001) which displayed no relationship with age.

**Conclusions:** Survival rates are encouraging. Protocols employed (traffic light-driven personalised care, protocolised early therapeutic anticoagulation based on D-dimer >1,000ng/mL and/or CRP>200 mg/L, personalised ventilatory strategies and antioxidants) are recommended to other units. Males are at greater risk of severe disease, most likely as the obligate SARS-CoV-2 receptor is encoded by the X-chromosome, and require especially close, and early attention.

## INTRODUCTION

Human infection due to the novel coronavirus SARS-CoV-2 (commonly referred to as COVID-19) was first reported at the end of 2019 [1]. The broad spectrum of disease severity ranges from asymptomatic and mild cases, to severe multiorgan failure and death. In early reports available from China [2], particular features were of multilobar viral pneumonia, frequent requirement for supplementary oxygen/ventilatory support, and very high transmission rates to healthcare workers. Subsequently affected countries have been able to benefit from the experience in Wuhan.

In the first 191 patients from Wuhan who had been discharged or had died by Jan 31, 2020, the crude death rate was 54/137 (39.4%) [2]. For intensive care, the initial experience in China was of 97% case fatalities following mechanical ventilation [2], with the development of adult respiratory distress syndrome (ARDS) in more than half of critically ill patients [2]. Secondary bacterial infection, and acute kidney, cardiac and liver failure were also commonly reported. Risk factors for in-hospital death were older age, higher Sequential Organ Failure Assessment (SOFA) score, and D-dimer greater than 750ng/mL on admission) [2].

Incorporation of these experiences into care pathways has led to reports of better, but still poor survival rates following hospitalisation. In the UK, the Intensive Care National Audit and Research Centre (ICNARC) recently reported 48.6% mortality in the first 5,139 patients with COVID-19 completing an Adult Intensive Care Units (AICU) admission [3]. This proportion was unchanged from earlier in the epidemic [4]. Similarly recent Italian and Washington State AICU reports indicate that for completed encounters, mortality was 61.2% and 67% mortality [5,6] respectively.

These data prompted us to report the outcomes from a single institution which had implemented novel procedural and management protocols, aiming to secure better survival rates for those infected by COVID-19 and requiring hospital admission.

## METHODS

### Institution Details

The Chelsea and Westminster NHS Foundation Trust comprises two hospitals and 12 community-based clinics in North-West London, and delivers acute care to a population of over 1.5 million people with socioeconomic descriptors detailed in [7]. The Accident and Emergency services provided at Chelsea and Westminster Hospital and West Middlesex University Hospital treat over 300,000 patients a year.

A new cloud database was created in Microsoft Azure with a near real-time pipeline from electronic patient records (EPR) and dedicated to COVID-19 reporting from an operational, nationally mandated and clinical perspective. The database was kept in a permanent development cycle to facilitate rapid additions and changes. As the volume of data in both patient numbers and extracted clinical data grew, a limited scale data mining exercise was undertaken to flag any presenting readings with unusually high correlation with certain outcomes, and shared with the first author (MPV). For management of COVID-19 cases, all healthcare staff were supported with personal protective equipment (PPE) appropriate to the level of aerosolisation risk, according to institutional guidance.

### Traffic light system integrating with electronic patient records

Bioinformatics presented in detail elsewhere (McCarthy et al, manuscript in preparation), delivers a near real-time decision support tool on the clinical dashboard for each electronic patient record. This is updated every 10 minutes from the first set of observations on emergency department admission. This was created for use with COVID-19 patients in March 2020, went live on 20^th^ March 2020, and is under continual development. Elements captured on all patients are described in the following section.

Symptoms captured included fever, dry cough, fatigue, dyspnea, diarrhoea, tachypnoea, thirst, loss of appetite. Observations included temperature, heart rate; blood pressure; respiratory rate; oxygen saturation (SpO_2_); fraction of inspired oxygen (FiO_2_, from 21% [room air] to 100%); oxygen delivery (nasal cannulae, face mask, continuous positive airway pressure (CPAP) and invasive ventilation).

Captured results of blood tests spanned full blood count including red and white cell counts (with differential for e.g. lymphocytes, neutrophils, monocytes, eosinophils); haemoglobin, haematocrit, and platelet count; coagulation screen including fibrinogen, prothromin time and activated partial thromboplasin time (APTT); D-dimer, and anti-factor Xa where appropriate; electrolytes (sodium, potassium), renal function (urea, creatinine), liver function (bilirubin, alkaline phosphatase, albumin, aspartate transaminase, alanine transaminase, gamma glutamyl transferase (GGT), lactate dehydrogenase (LDH)), iron studies (serum iron, transferrin saturation index, ferritin); and additional markers of the acute phase response (C-reactive protein, fibrinogen).

Elements of therapeutics were also captured spanning ventilatory support including FiO_2_ (as above), circulatory support including noradrenaline, and renal replacement therapies With evolving clinical experience, addditional markers of disease severity were included on the dashboard including procalcitonin (PCT).

In the first phase, a daily report was issued on all COVID-19 positive patients in the Trust, and sent to an experienced critical care clinician (MPV) for review. Results were interpreted and fed back to the clinical teams to modify pathways, particularly regarding anticoagulation, radiological imaging results; trial of continuous positive pressure ventilation, and early transfer to AICU for additional interventions:

### Anticoagulation

For prophylaxis of venous thromboemboli (VTE), recognising the data from [2], the usual institutional prophylactic dose of enoxaparin (clexane) 40mg once daily was increased to an intermediate dose of 40mg twice daily for all admitted COVID-19 positive patients. Dashboard markers of D-dimer >1,000ng/mL, heart rate <50 or >100min^-1^ and/or CRP >200mg/L resulted in urgent feedback from the AICU Consultant to the clinical team of a high likelihood of thromboembolic disease. These patients underwent a computerised tomographic pulmonary angiogram (CTPA) to screen for an overt pulmonary embolus (PE) and a non-contrast computerised tomogram of the brain (CTB) using established institutional protocols, to exclude intracranial haemorrhage or an acute cerebral infarct, prior to initiating systemic anticoagulation. In the absence of visible thrombus on CTPA, a microvascular thrombosis was assumed as previously reported [2], and therapeutic anticoagulation usually commenced. However, if CTB identified an acute cerebral infarct which could be at risk of haemorrhagic transformation, anticoagulation was maintained at prophylactic dose.

### Ventilation and oxygenation assessments

Dashboard markers of a high respiratory rate (> 35/min^-1^); SpO_2_ <92%, inspired fractional partial pressure of oxygen (FiO_2_) >60% or deterioration of haemodynamic function were integrated by the AICU Consultant in the emergency department. Patients meeting the criteria received a 30 minute trial of CPAP in the emergency department. If the respiratory rate fell by 30% or more, CPAP was continued, otherwise patients were intubated and ventilated inmediately by a dedicated intubating team and transferred to AICU for invasive ventilation.

### Adult Respiratory Distress Syndrome (ARDS) /Secondary Infection

This pattern was defined in ventilated patients with dashboard markers of high temperature (>38°C), FiO_2_ >60% with SpO_2_ <94%, positive end expiratory pressure (PEEP) of more than 10mmHg, CRP >250mg/L, requirements for noradrenaline to support the circulation, and a rising serum fibrinogen >5g/L. In this pattern, platelets were also reduced. Patients were noted to have a poor response to high FiO_2_, relatively good response to ventilation with prone positioning, but patients were noted to not tolerate the previously recommended conservative fluid balance for (COVID-19) ARDS [8], with renal function deterioration.

### Hyperinflammatory/’cytokine storm’ indices

This pattern was observed with sudden haemodynamic collapse after relative stability. Dashboard markers of temperature >39°C, diastolic blood pressure <35mmHg, CRP >250mg/L and ferritin >1,000µg/L were used to define patients at medium and high risk of a hyperinflammatory response. Patients meeting the criteria were monitored continuously and fluid management adjusted to replace insensible losses (1ml.kg^-1^.h^-1^ for every degree above 38°C). If there was no response to initial fluid management and diastolic blood pressure remained <35mmHg, or oxygen requirement increased, then the patients were referred to the AICU for further management.

### COVID-19 Critical Care Protocols

Patients who did not improve following a trial of CPAP, or meeting other criteria for AICU referral, were transferred to intensive care units at the two hospitals. There, they were intubated, ventilated using volume/pressure support, and managed according to conventional protocols, but with four additional modifications: 1) Ventilatory settings aimed for a target SpO_2_ 92 % and generally low initial PEEP of 8cm water (H_2_O), as lung compliance was observed to be normal/high during the early phase of the disease process. 2) Ventilatory position employed very early prone ventilation, within first 4 hours of admission to the AICU. 3) For fluid balance, an even balance was targeted, with the use of diuretics where appropriate, unless there was a positive passive leg raise which suggested low intravascular volume. A negative balance was targeted in patients with features suggestive of ARDS i.e. low lung compliance and presence of B-lines on lung ultrasound. 4) Ascorbic acid 1mg bd was given given enterally or intravenously to all AICU patients by protocol to reduce oxidant stress in the setting of shock/microvascular shutdown.

### Data Analyses

Selected dashboard data including the real-time flags for VTE, ARDS, and cytokine storm were downloaded. Notably, for calculation of the population age deciles, all cases with the boundary age were assigned to the lower or upper decile which contained most of the values (none were equally distributed between two deciles).

Separately, to explore genotypic predisposition, DNA variants in the *ACE2* gene were downloaded from the Genome Aggregation Consortium (gnomAD) [9]. This had mapped largely non-overlapping datasets of 213,158 exomes and genomes from 111,454 males and 101,704 females to human genome builds GRCh37/hg19 [10] (version 2) and GRCh37/hg19 [11] (version 3) [9].

Statistical analyses were performed in Stata IC version 15 (Statacorp, Texas) to generate descriptive statistics, graphical illustrations, to compare datasets, and to perform logistic and linear multiple regression. Categorical comparisons were performed using χ^2^. Continuous data comparisons were performed using non-parametric tests suitable for all distributions (Mann Whitney U test). Selected data were also illustrated using Microsoft Excel, Microsoft Powerpoint, and GraphPad Prism 8.1.1 for Windows, GraphPad Software, San Diego, California USA, www. graphpad.com.

## RESULTS

### Population demographics

By 22^nd^ April 2020, 923 patients admitted to the two hospitals had tested positive for COVID-19 (*Figure 1*). The median age of patients was 67ys (interquartile range, IQR 53-80 ys), and comprised 569 males (61.7%), and 354 females (38.4%). The population comprised approximately equal proportions of White (346 recorded), and Black and Minority Ethnic (326 recorded). Compared to other reported cohorts [12], the population were already in an advanced state of disease at the time of admission (*Supplementary Table* 1). In particular, the median CRP was 102.1mg/L [interquartile range, IQR 44.9-179]; median D-dimer 1920ng/mL [IQR 1070-3802], and ferritin 566µg/L [IQR 261-1208].

**Figure 1:**
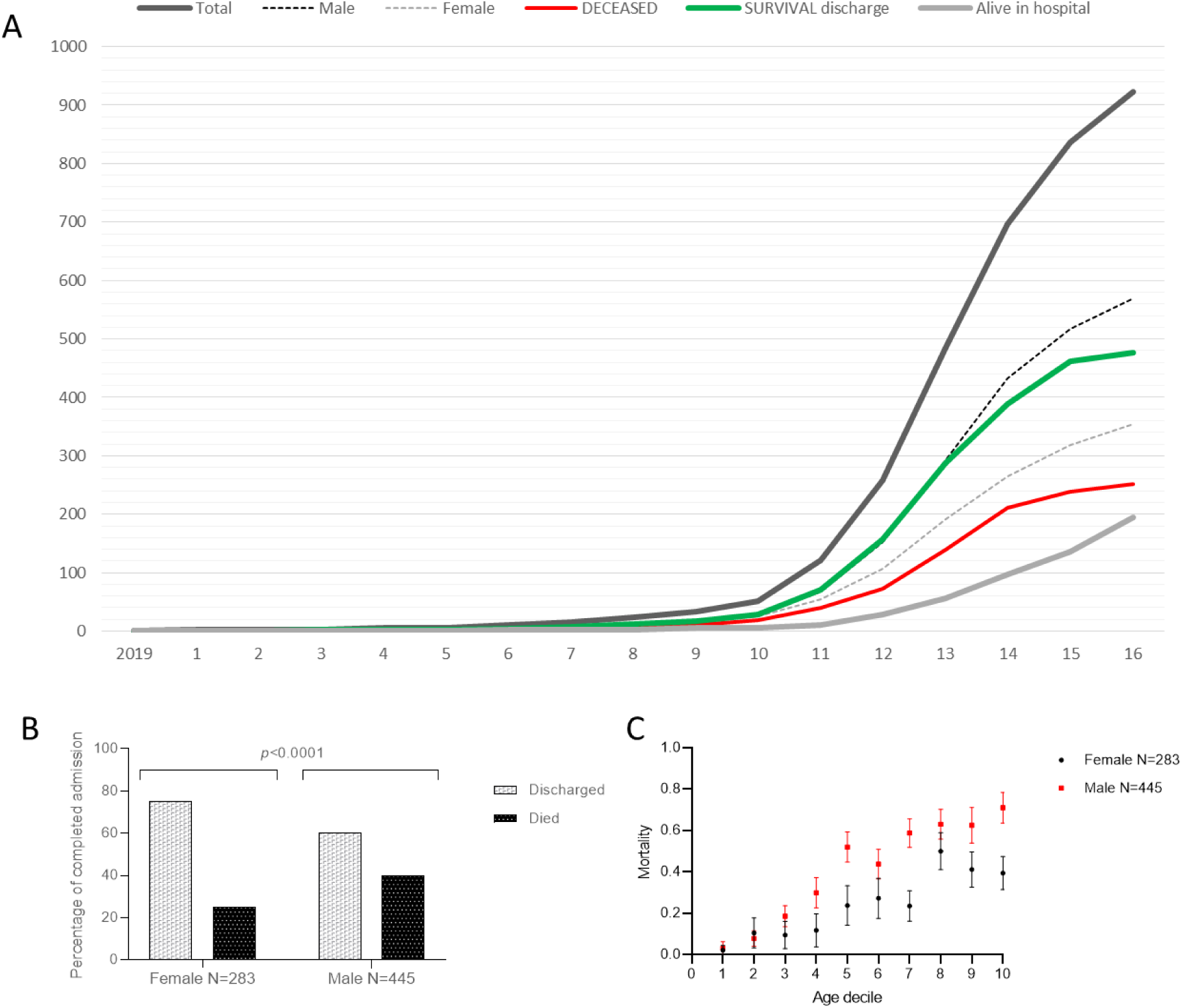
**COVID-19 cases in institution**. **A:** Cumulative cases across the first 16 weeks of 2020 until 22 April 2020. Total COVID-19 cases are plotted in thick black, males by black dotted line, females by grey dotted line, cases who survived to discharge in green, cases who died in red, and cases still in hospital by thick grey line. **B)** Comparison of male and female outcomes across all ages, using cases restricted to known outcomes (*p* value calculated by χ^2^ with 1 degree of freedom). **C)** Comparison of male and female outcomes across age deciles (age range 0-106 years, decile boundaries at 39, 50, 56, 61, 67, 73, 77, 82 and 87 years). The *p* value was <0.001 calculated by logistic regression using absolute age (years), or age deciles as illustrated.

On data download 23/04/2020, 195 patients were still in hospital, and 139 had been admitted to the Adult Intensive Care Unit (AICU). Patients median length of stay was 6 days [IQR 3, 10] without AICU (N=449), 11 days [IQR 7.5, 20.5] if admitted to AICU including a median stay of 3.5 days [IQR 2,5, 7.1] on AICU (N=28). For patients still in hospital, the median length of stay was 9 days [IQR 5, 17] if not admitted to AICU (N=108), 19 [IQR 15, 27] if admitted to AICU including a median stay of 15.5 days [IQR 7.6, 21.3] on AICU (N=45).

### Case Fatality rates higher in males

Whether evaluating all cases, or cases restricted to known outcomes only, males displayed worse outcomes than females (χ^2^ p<0.0001, *Figure 1B*). The overall case fatality rate estimated for males and females based on the 728 ‘completed’ cases was 25.1% [95% CI 20.0, 30.0] in females, 40.4% [95% CI 35.9, 45.0] in males. The excess male mortality was seen in all age groups (*Figure 1C*).

One hundred and thirty-nine patients were admitted to Adult Intensive Care (AICU). AICU admissions were younger than non AICU patients (mean age 56ys [95% CI 54, 58ys] versus 66ys [95% CI 65, 68ys]. AICU patients were more likely to be males (N=102/139, 73.4%), more likely to be from an ethnic minority (60/99 recorded, 60.6%), and sicker in all parameters tested (data not shown). Currently 94/139 patients are still alive, compared to 578/784 ward admissions (AICU survival 67.6% [95% CI 60, 76], general ward 73.7% [95% CI 71, 77%]). Again, even with the small AICU numbers, the striking finding was an excess of male deaths: Of 45 AICU deaths, 39 (86.7%) were in males. At the time of manuscript submission, the overall death rate amongst AICU admissions was 32.4% [95% CI 24.5, 40.2]. Amongst males it was 38.2% [95% CI 28.6, 47.8], and amongst females 16.2% [95% CI 3.8, 28.7], with no trends for differences based on ethnicity.

### Traffic Lights Real Time Monitoring from Emergency admission

From initial admission in the Emergency units, patient data were autopopulated into the hospital dashboard, and updated every 10 minutes. Real time-reports were sent to an experienced critical care clinician (MPV) for review. Results were interpreted and fed back to the clinical teams. Patterns identified allowed rapid modification of treatment pathways that were individualised to each patient, but particularly regarded anticoagulation, imaging, trial of continuous positive pressure ventilation, and early transfer to AICU for additional interventions (*Figure* 2).

**Figure 2:**
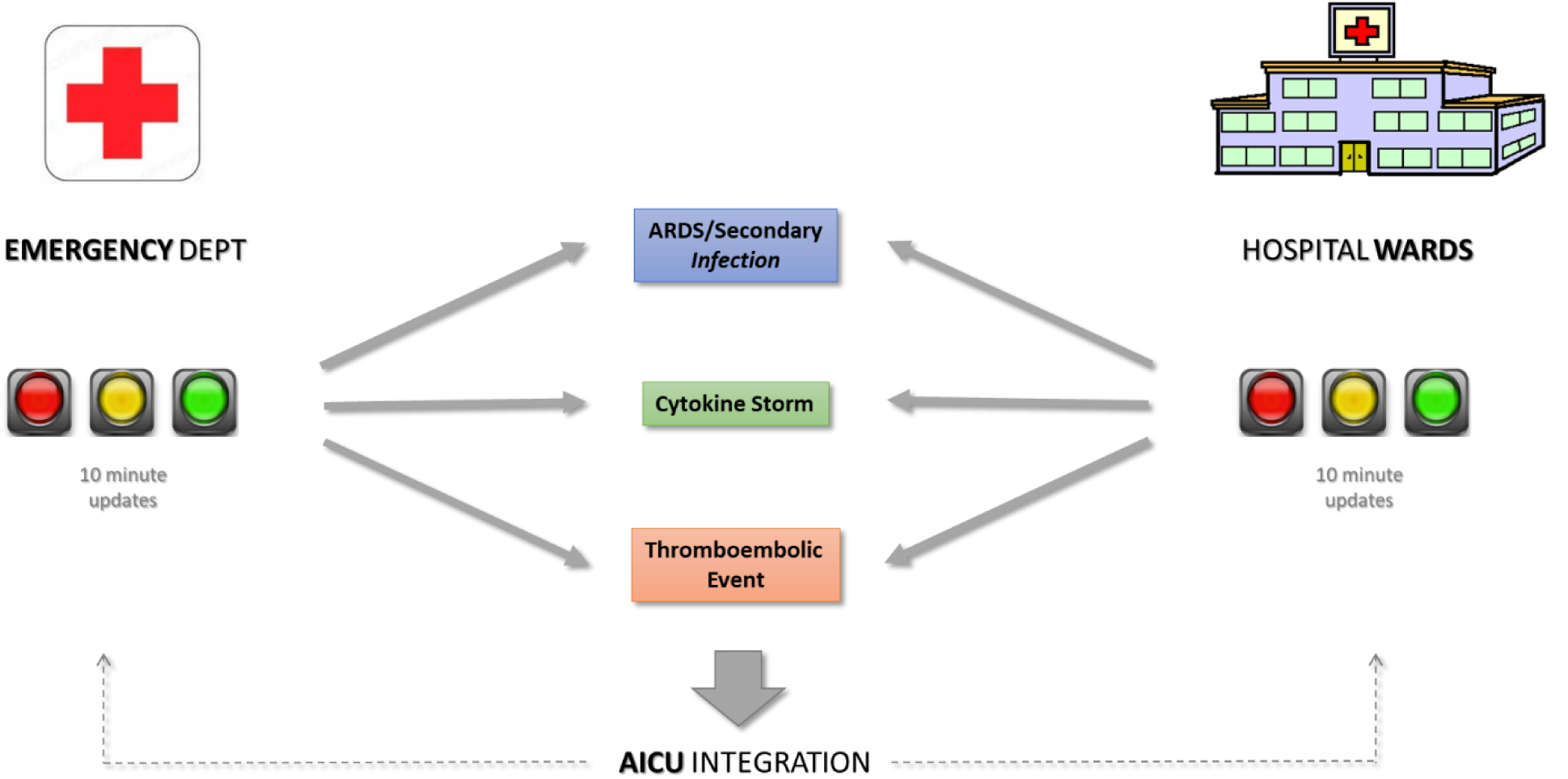
**Real time decision support tool** highlighting the 3 patterns of disease that were recognised early in the outbreak to be causing mortality. In pattern 1, the disease remained single-organ, usually respiratory but progressed from a viral pneumonia to a severe adult respiratory disress syndrome (ARDS) +/- superimposed bacterial pneumonia. Pattern 2 was characterised by circulatory collapse (cytokine storm), and pattern 3 by thromboembolic disease.

### Patterns of critical disease phenotypes in COVID-19 Patients

Amongst the 161/923 (17.4%) of admissions with traffic light records of normal temperature, diastolic blood pressure, respiratory rate and low FiO_2_, only 18 had a ferritin <300 µg/L, CRP <50 mg/L and D-dimer <3,000 ng/mL. In other words, at presentation, almost all patients were sicker than might have been expected based on standard observations. The 18 comprised 11 males and 7 females, median age 76 ys [IQR 66, 84ys]. None required AICU care, their median hospital length of stay was 10.5 days [IQR 4, 20 days], and overall 12 are already discharged, two (11.1%) died, with 4 still in hospital.

Nine-hundred and five patients (98.1%) had markers of severe physiological derangement. The proportions in the 3 distinctive patterns of critically ill patients were not equal and are illustrated in *Figure 3*. All groups had an excess of males, noting males had represented 61.7% of all admissions (85% for ARDS-flagged cases, 82% for cytokine storm, 66% for all pulmonary, and 55% for thromboembolic-flagged cases).

**Figure 3.**
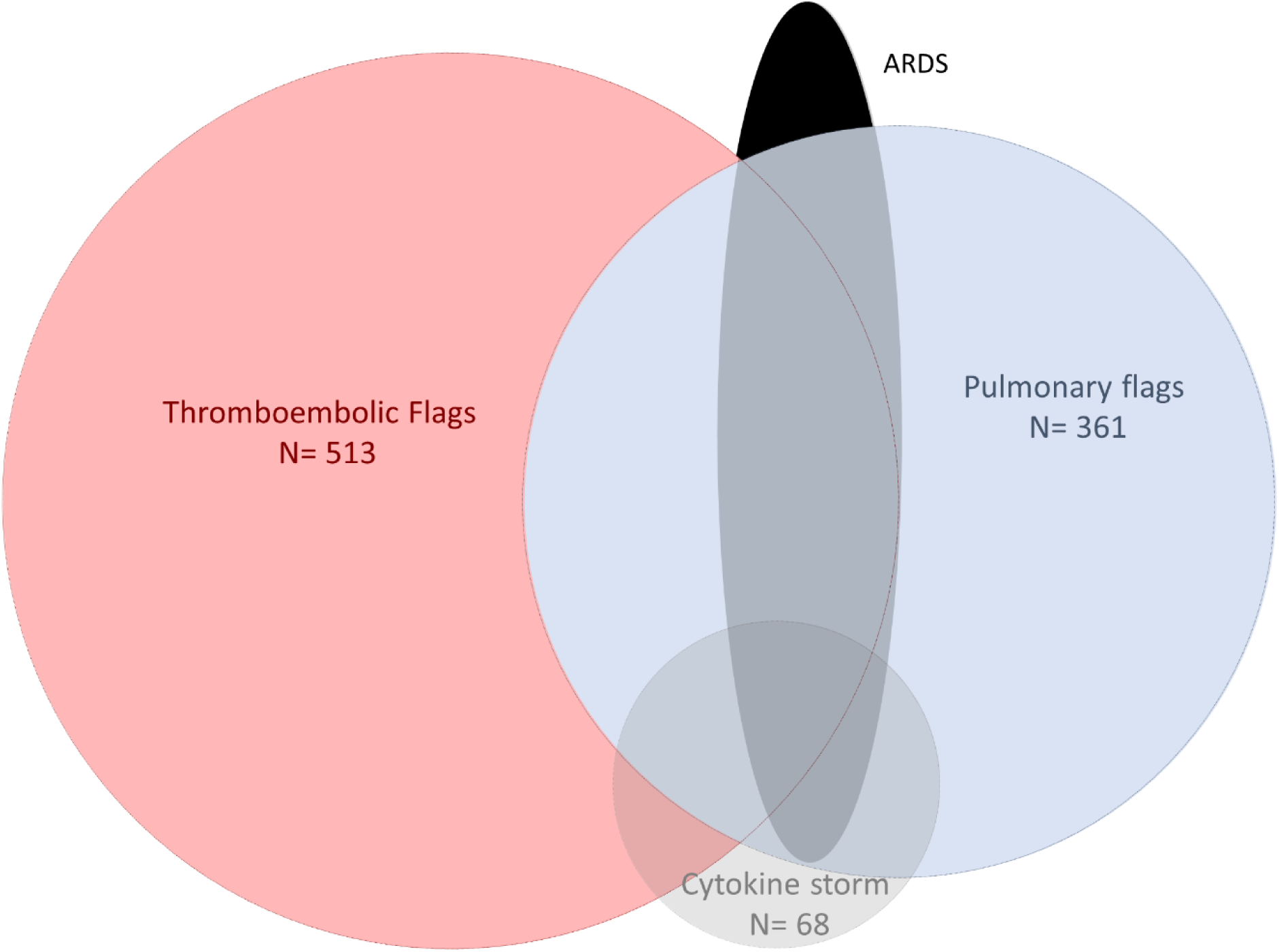
**The distinctive patterns of critically ill patients** Relative proportions of patients falling into dashboard-flagged categories are drawn to scale by area and overlaps of respective circles. The ARDS component is not fully to scale since due to delays in set up, only 122 patients had the procalcitonin results and therefore the total number was estimated for a population of 923 cases.

### Dashboard biomarkers highly abnormal in males, but no association with increasing age

Noting that males were more likely to die, we examined differences in biomarkers between males and females. Males were younger than females (median 65ys [IQR 54, 78], compared to median 72ys [IQR 52, 82] for females, p = 0.037). Despite this, all key dashboard variables tended to be more abnormal in males. Notably, the association was significant to *p* <0.0001 for CRP, ferritin, and creatinine, and p<0.001 for respiratory rate and diastolic blood pressure (*Supplementary Table* 1). CRP was selected for illustration: as shown in *Figure 4*, the increase in CRP in males was sustained across all age groups. Similar findings were observed for ferritin, illustrated in *Supplementary Figure 1*.

**Figure 4:**
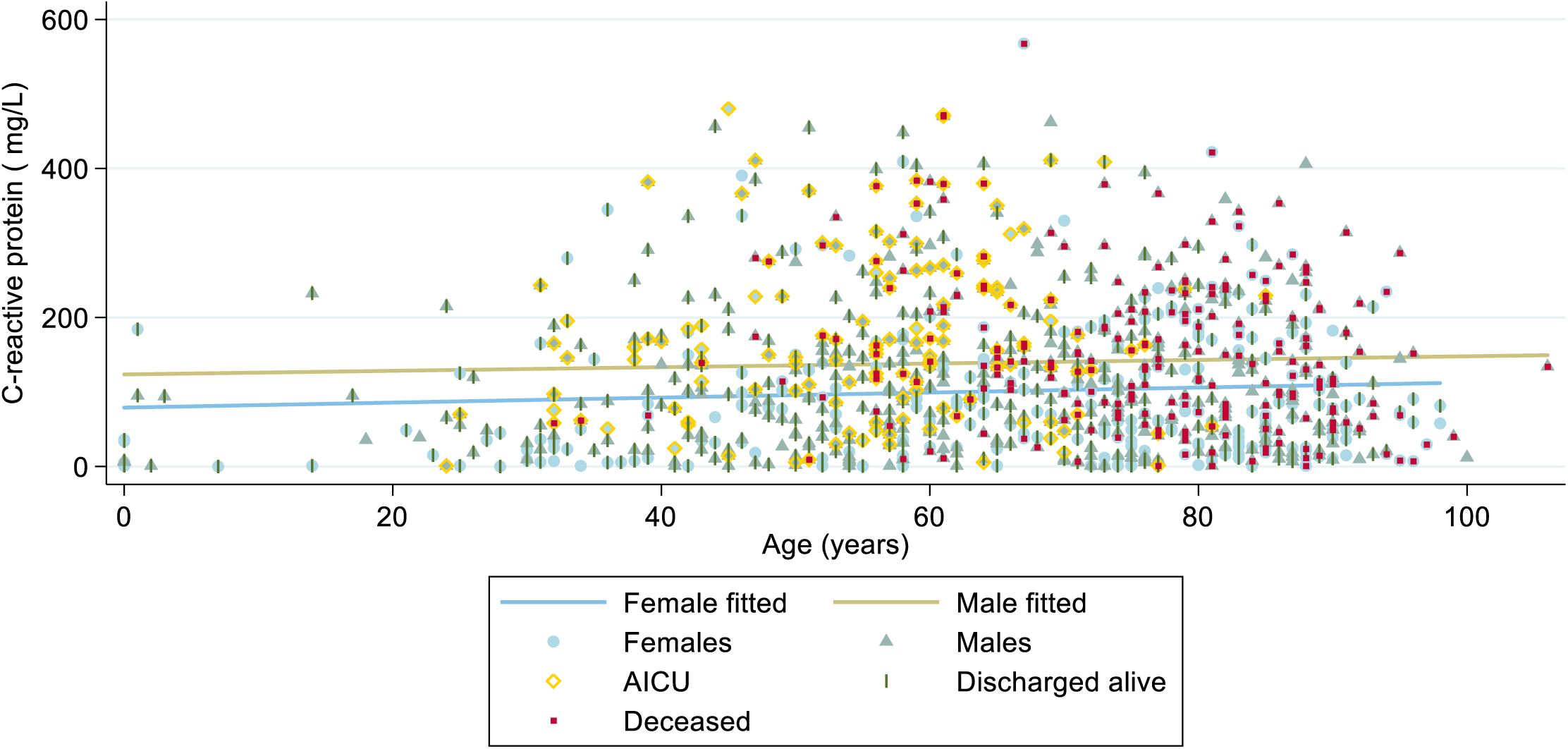
**CRP values across all age groups by sex and outcome**. Individual values and regression lines are plotted separately for all COVID-positive males (green triangles) and all females (blue circles) admitted to the Chelsea & Westminster NHS Foundation Trust by 22nd April 2020. Superimposed on their symbols are annotations indicating whether the patient was on AICU (yellow ring), and for completed encounters, whether the patient was discharged alive (green pipe), or died (red square). For further illustration with regression line confidence intervals, see *supplementary Figure 1A*

As indicated in *Figure 4*, there was minimal association between age and CRP in either sex. Furthermore, while there were physiological parameters that did differ according to the age of the patient, in the COVID-19 cohort there was no age-relationship with the core critical markers of CRP or ferritin, and only a marginal increase in male D-dimer values with age (*Supplementary Table 1, Supplementary Figure 1*).

Notably, all relationships with gender persisted after adjustment for age (*Supplementary Table 1*).

### Male gender may predispose to COVID-19 through unopposed ACE2 sequence on X-chromosome

A plausible biological reason for why males are at heightened risk of severe pathophysiological derangements and death following COVID-19 infection is provided by the fact that protein which SARS-CoV-2 uses as a “receptor”, is encoded by the *ACE2* gene on the X chromosome. The protein is better recognised as angiotensin I receptor 2 (ACE2). Males with their [46, XY] karyotype have a single X chromosome, and, as evident by X-linked disorders such as haemophilia, have more severe consequences from single detrimental alleles than females.

We estimated the degree of relative risk for having a COVID-susceptibility allele (e.g. one that increased *ACE2* expression) by examining variants in the gnomAD databases: These had mapped datasets from 111,454 males and 101,704 females to human genome builds GRCh37/hg19 [10] (version 2 and GRCh37/hg19 [11] (version 3) [9].

In the *ACE2* exons that encode the protein, 23,517 variants were the unopposed *ACE2* sequence in males compared to 6,024 in females. The hemizygosity rate in males (21.1 per 100) compared to homozygosity rate in females (5.9 per 100) indicates that males are approximately 4-times as likely to have no “normal” *ACE2* sequence (*Figure 5*).

**Figure 5:**
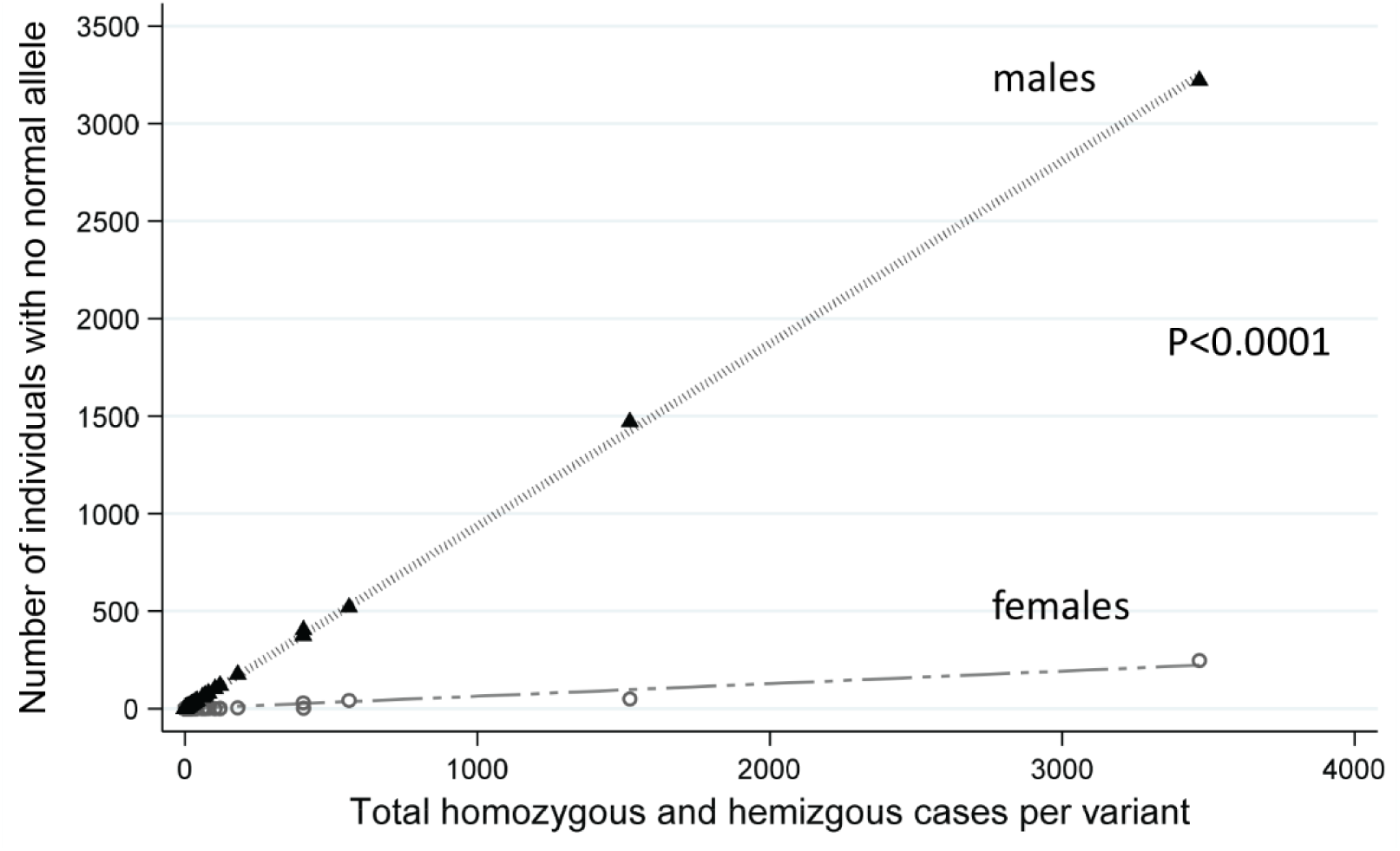
**Number of individuals with no normal ACE2 sequence**. The x axis plots the total number of cases with no normal allele at particular DNA nucleotides, and the y axis indicates the number of males (triangles) or females (open circles). While the male bias pattern is observed at all allele frequencies, it is most clearly illustrated focussing on more common allelic variants as illustrated.

## DISCUSSION

We defined three critical disease groups by using a real-time, traffic lights dashboard that enabled rapid adjustment of management protocols. There was minimal evidence that age was associated with disease severity, but males were significantly more likely to have severe disease biomarkers, and to die, including following admission to AICU. We have also demonstrated from an acute hospital setting admitting patients in an advanced stage of their illness, that overall death rates of 20.5% [95% CI 16.0, 25.0%] on general wards, and 16.2% [95% CI 3.8, 28.7] for AICU admissions can be achieved for females.

In the final stages of manuscript preparation a study of 5,700 patients admitted in New York has published similarly low mortality, though notably that cohort included a higher proportion with milder disease on admission as evidenced by admission CRP and D-dimer values [12]: the New York interquartile ranges [12] did not overlap with the interquartile range of the current cohort (Supplementary Table 1), instead representing the lowest 160 (17.3%) and 149 (16.1%) for current cohort CRP and D-dimer respectively.

Given the current cohort were already at an advanced disease state on presentation, one reason for their generally good survival rates is likely to be the real time Clinical Decision Support tool that enabled rapid escalation of treatments, particularly relevant since the commonest critical scenario was thromboembolic disease for which anticoagulation is available: It was recognised early that the thrombotic process was most amenable to preventative and early treatment strategies, and more recent data have emphasised this: Overall thrombotic complication rates in intensive care patients are now reported as exceeding 30% [13–15]. Recent data from severely affected patients suggest the coagulopathy is not consistent with acute disseminated intravascular coagulation (DIC) and instead supports hypercoagulability together with a severe inflammatory state [16]. Additionally, autopsy series published in recent weeks highlight particularly the thrombosis of small vessels [17,18], and multiple groupings are now advising on how best to manage anticoagulation in COVID-positive patients [19-23]. Recognising evidence of microvascular injury [17,18], it is plausible that early, stratified intermediate prophylactic or full therapeutic anticoagulation, where appropriate, with heparin may have modified the disease process and led to improved outomes in our protocolised case series compared to all series [4–7; 12], and low overall admission mortality rates for a cohort already at an advanced stage of disease. Furthermore, independent of heparin’s anti-coagulant properties, it may exhibit important anti-viral properties. Lang and colleagues [24] have shown that heparin inhibits SARS-CoV infection *in vitro* and it has been shown to interact with the spike protein of SARS-CoV-2 causing conformational change, which may inhibit interaction with the ACE2 receptor and thereby block host cell entry[25].

The addition of an antioxidant in the form of ascorbic acid, for all patients admitted to hospital, could also have contributed to lower mortality in our case series. Although not shown to improve outcome in small sepsis [26] and ARDS [27] studies, there is growing interest in the use of ascorbic acid in sepsis [28]. Ascorbic acid has been shown to shorten the duration of the common cold and reduce hospital stay in influenza-A associated pneumonia. [29] *In vitro* studies have demonstrated that ascorbic acid enhances the proliferation, differentiation and function of lymphocytes, and stimulates interferon release,[30] which could enhance viral clearance.

There is substantial media discussion that older individuals, and people from ethnic minorities, are more at risk from COVID-19. While we cannot discount such signals, the evidence was not strong in the current cohort. Instead the most striking feature was the male predominance- in ward admissions, in admission to AICU, for biomarkers of severe disease (especially CRP and ferritin that displayed no relationships with age), and for deaths. We are not the first to report male susceptibility to severe COVID-19 [1–7], but the evidence for susceptibility to all aspects, across all age-groups, is noteworthy. The genomic evidence from *ACE2* adds to the compelling clinical data that emphasise the importance of increasing targeted and personalised care to males who are biologically more at risk of severe complications if infected by COVID-19. The presented biomarker data and excess mortality seen on AICU imply however, that even with the accelerated in-hospital care provided by the traffic light system, as a group, males are not benefitting as much as females. The nature of putative X-chromosome variants that modify the expression of the viral receptor are yet to be determined, thus for now we suggest that all males should be currently considered to be at higher risk.

In conclusion we define the three major patterns of COVID-19 mortality, present a traffic light system that enables early anticoagulation and other accelerated care pathways, and report encouraging survival rates in a hospitalised cohort admitted with advanced disease. We found no evidence that age was associated with biomarkers of disease severity- in contrast these emphasised the males at risk. We suggest the presented protocols are adapted more widely. Earlier presentation of males to hospital is also encouraged, in efforts to prevent the pathophysiological deterioration to which they are biologically susceptible.

## Data Availability

Fully anonymised data will be available post peer review publication on reasonable request, in accordance with institutional protocols.

## Authors’ Contributions

The study was conceived and overseen by MPV. Literature searches were performed by MPV, CLS MH and RD. The traffic light bioinformatics system was designed and implemented by AH, AB and AM. The clinical study design was by MPV. Data analysis for real-time feedback was performed by MPV and AM. Real time analytic data application was performed by MPV, MH, SS, LC, AS, RD, CL, MP, AG, JA, TP and RTK. Data analysis was performed by CLS, and data interpretation was performed by MPV and CLS. Figures were generated by CLS. The manuscript was written by CLS, and revised by MPV, MH, RD, AM, and RTK. All authors reviewed and approved the final manuscript.

## Acknowledgement

The authors wish to thank the Chelsea & Westminster NHS Foundation Trust personnel and especially the Adult Intensive Care Unit nurses, for the delivery of personalised care to all patients admitted with COVID-19 infection. The authors would also wish to thank the CW Plus charity for invaluable support throughout the COVID-19 outbreak. A special thanks to Dr Simon M Greenfield for revising the manuscript for grammar and syntax

## Ethics Committee Approval

This work was approved by the Chelsea & Westminster NHS Foundation Trust Clinical Governance team and the Data Protection Officer, Head of Information Governance. As we report on routinely collected non-identifiable clinical audit data, no ethical approval was required under the UK policy framework for health and social care.

## Data Availability

Fully anonymised data will be available post peer review publication on reasonable request, in accordance with institutional protocols above.

---

Early detection of severe COVID-19 disease patterns define near real-time personalised care, bioseverity in males, and decelerating mortality rates.

Marcela P. Vizcaychipi MD PhD FRCA*^1,2^, Claire L Shovlin PhD FRCP^A3,4^, Michelle Hayes MD FRCA^1^, Suveer Singh PhD FRCP^1,6^, Linsey Christie FRCA’, Alice Sisson FRCA^1^, Roger Davies FRCA^1^, Christopher Lockie FRCA^1^, Alice Howard^5^, Alexander Brown ^5^, Alex McCarthy^5^, Monica Popescu FRCA^1^, Amandeep Gupta FRCA^1^, James Armstrong FRCA^1^, Hisham Said FRCA^1^, Timothy Peters FRCA^1^, Richard T Keays MD FRCA^1,2^, ChelWest COVID-19 Consortium!

DATA SUPPLEMENT

Supplementary Table 1: Comparison of dashboard indices on admission, by sex………. 22

Supplementary Figure 1: Comparison of key dashboard indices on admission by age sex and outcome………. 23

Supplementary Table 1

**Supplementary Table 1:**
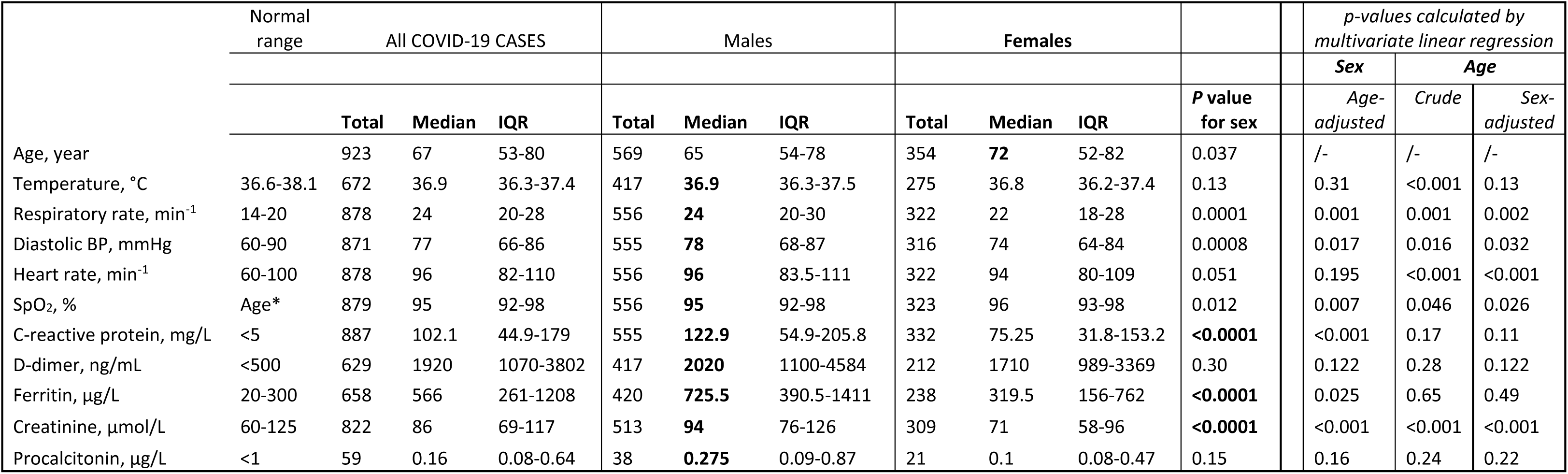
**Comparison of dashboard indices on admission, by sex**. Bold face indicates the more abnormal variable between the sexes, and for *p*-values, those meeting p<0.0001. Females were older, there was no difference in the percentage of males and females by ethnicity, but all dashboard variables tended to be more abnormal in males, particularly CRP (illustrated in main manuscript *Figure* 4), ferritin, creatinine (*p*-values <0.0001); respiratory rate and diastolic blood pressure (p<0.0001). BP, blood pressure, SpO_2_, oxygen saturation, IQR, interquartile range. P values for sex calculated by Mann Whitney rank sum. * Age dependent, SpO_2_ normal low 92% (for older patients), normal high 98.5%. The final columns indicates the *p*-values following exploratory multivariate regression analyses (regression coefficients not indicated for clarity).

Supplementary Figure 1

**Supplementary Figure 1.**
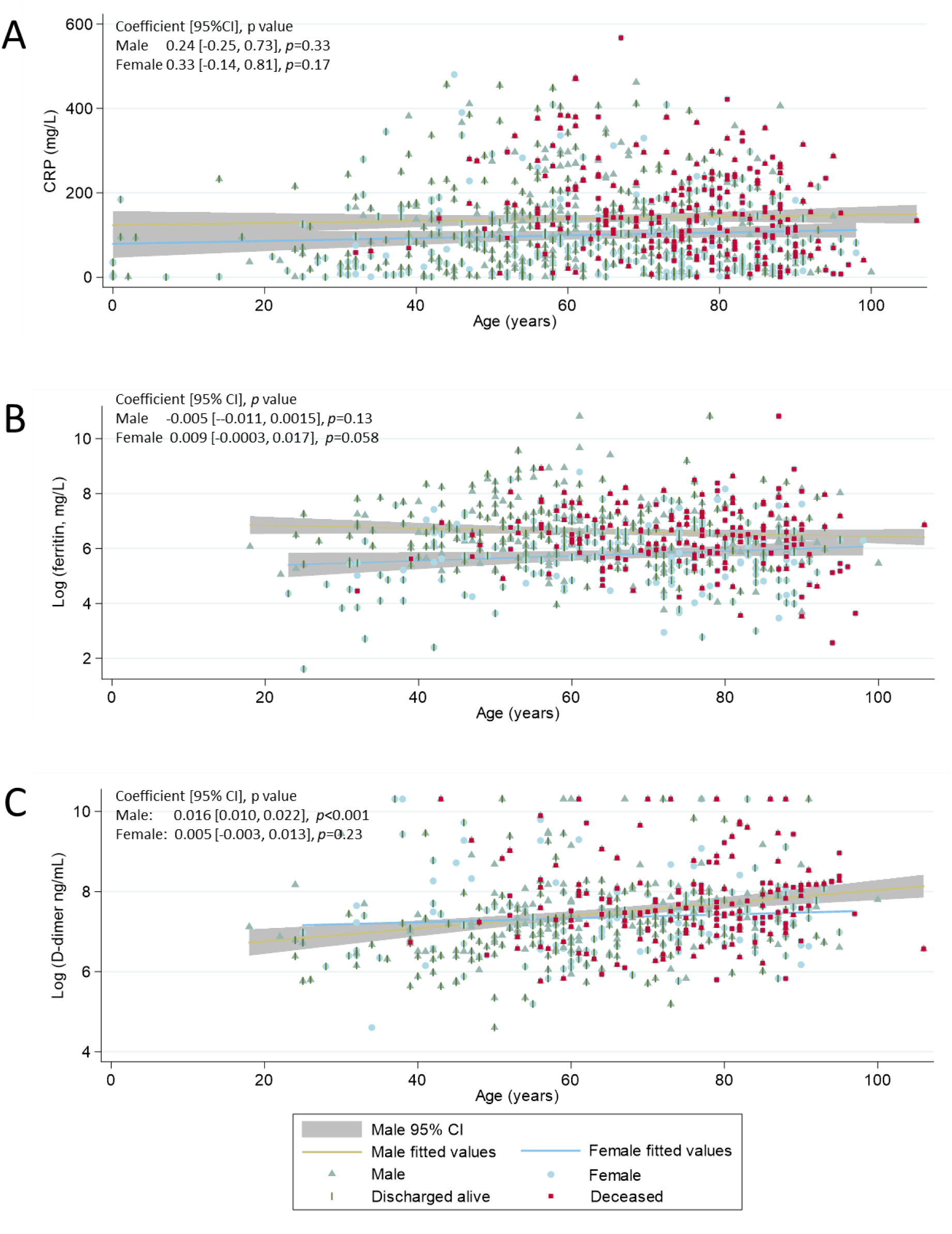
**Comparison of key dashboard indices on admission by age sex and outcome**. Individual values are plotted for males (green triangles/ khaki regression line), and females (blue circles/blue regression line). Superimposed for completed encounters, are whether the patient was discharged alive (green pipe), or died (red square). A) Admission CRP values including 95% confidence intervals [CI] for males and females, and illustrating higher values in males that becomes less pronounced at older age as female ferritin tends to increase. C) Natural log-transformed D-dimer. Note there was again no relationship with age for females (95% CI not shown for clarity), but that D-dimer increased with age in males. However, the regression coefficient for ln (D-dimer) predicted an averaged increase in the order of 10ng/mL per decade which we considered unlikely to have a bearing on the values under discussion.

